# Number of tests required to flatten the curve of coronavirus disease-2019

**DOI:** 10.1101/2020.12.26.20248818

**Authors:** Seogwon Hwang, Jong-Hoon Kim, Young June Choe, Dong-hyun Oh

## Abstract

We developed a mathematical model to quantify the number of tests required to stop the spread of coronavirus disease 2019 (COVID-19). Our model analyses performed using the data from the U.S. suggest that the infection coefficient increases by approximately 47% upon relaxing the lockdown policy. To offset the effect of lockdown relaxation, the number of tests should increase by 2.25 times, corresponding to approximately 280,000–360,000 tests per day in April 2020.

---

The lack of vaccines has led to the adoption of non-pharmaceutical interventions, such as case isolation and contact tracing, as public health measures for containing the transmission of coronavirus disease 2019 (COVID-19) (1). However, syndromic surveillance devoted to identifying symptomatic people will unlikely be sufficient for stopping transmissions because a substantial number of transmissions are attributed to asymptomatic cases (2, 3). Therefore, identifying asymptomatic transmitters, who may escape syndromic surveillance, is critical to controlling the spread of COVID-19. Here, we propose a mathematical model that can provide insights on the number of tests required to identify infectious people and stop them from spreading COVID-19.

## Assumptions and Modeling

A substantial number of people infected with SARS-CoV-2 do not develop symptoms; thus, it would be significantly challenging to quantify the total number of infected people in real time. Therefore, rather than using classic SIR and SEIR models, we developed a mathematical model that includes the number of tests and confirmed cases, whose quantification and comparison with the data is more straightforward.

A well-known epidemic model can be modified as follows:

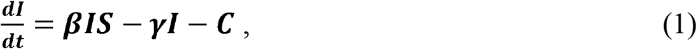

where *I* denotes the proportion of infected cases and *S* the proportion of susceptible population. The term *γ* denotes a coefficient that determines the rate at which the infected individuals recover without being confirmed, as they have mild or no symptoms. Additionally, *C* denotes the proportion of infected people detected through testing and isolated such that they cease to be infectious per unit time.

Cases are confirmed by testing people suspected of COVID-19 infection. Assuming that symptoms are exponentially distributed for the people who are suspected of infection and tested, we obtain the following relationship between *C* and the proportion of population tested per unit time *T*:

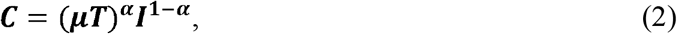

where *μ* and *α* denote appropriate model parameters. For detailed derivations, refer to the Supplementary Material. Equation (2) is of the form of the Cobb–Douglas function, which is frequently used in economics.

Substituting equation (2) into equation (1), we obtain the following:

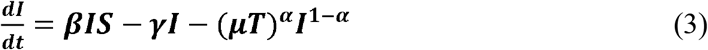

If the epidemic is in its early phase (*i*.*e*., *S*= 1) and equation (3) is equal to zero, the minimum number of tests, *T*^*^, required to suppress the spread of the disease can be obtained by substituting *I* with *C*, as follows:

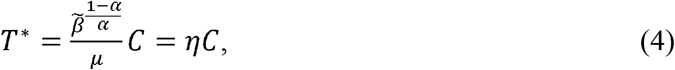

where 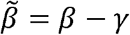.

Equation (4) shows that the number of confirmed cases provides information regarding the minimum number of tests required for preventing the spread of the disease (i.e., 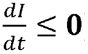). To stop the spread of COVID-19, the number of tests should be at least times the number of confirmed cases.

## Model fitting

We fit the model to the data using the following equation, which was derived from equations (2) and (3) (see Supplementary Material for details):

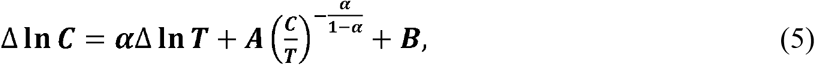

where 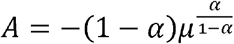 and 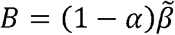. Additionally, Δ **In*C*** denotes the daily rate of change in the number of confirmed cases per population ; Δ **In *T*** denotes the daily rate of change in the number of people who are tested for COVID-19 per population; A and B are constants. In equation (5), *B* is related to the rate of infection and is affected by social distancing or lockdown policies.

## Results

The results obtained using the estimation strategy described in Supplementary Material are summarized in Table 1. For all the dates, the parameter estimates were statistically significant, except the coefficient estimates that corresponded to the dummy variables (*ld*) of the lockdown order for the early dates. However, the effect of the lockdown order became apparent from April 11, 2020, with statistical significance. The signs of all the estimates were in line with our expectations. The magnitudes of the change in the parameter estimates were negligible. Notably, *α, A, B*, and *ld* were estimated to be approximately 0.43, –0.08, 0.385, and –0.12, respectively. However, the parameter estimates may not significantly vary unless unanticipated events occur. Figure 1 shows the estimation results of *η* and *T*^*^ obtained using the above-mentioned estimation results. The red and blue lines indicate *η* and *T*^*^, respectively. The value of *η* ranges between 4.667 and 5.687, indicating that even if the lockdown order is enforced, the number of tests conducted daily must be at least approximately 5.7 times the number of daily confirmed cases, to prevent the spread. This means that the number of tests required to prevent the spread should have been, on average, more than approximately 150,000, in April.

**Table 1.** Estimation results.

|  | ~20200405 | ~20200406 | ~20200407 | ~20200408 | ~20200409 | ~20200410 |
| --- | --- | --- | --- | --- | --- | --- |
| $\alpha$ | 0.4274<br>(0.059)<sup>a</sup> | 0.4281<br>(0.058)<sup>a</sup> | 0.4286<br>(0.057)<sup>a</sup> | 0.4284<br>(0.056)<sup>a</sup> | 0.4290<br>(0.055)<sup>a</sup> | 0.4289<br>(0.054)<sup>a</sup> |
| A | -0.0808<br>(0.046)<sup>b</sup> | -0.0804<br>(0.045)<sup>b</sup> | -0.0802<br>(0.044)<sup>b</sup> | -0.0802<br>(0.043)<sup>b</sup> | -0.0800<br>(0.042)<sup>b</sup> | -0.0799<br>(0.042)<sup>b</sup> |
| B | 0.3870<br>(0.129)<sup>a</sup> | 0.3864<br>(0.126)<sup>a</sup> | 0.3863<br>(0.124)<sup>a</sup> | 0.3861<br>(0.123)<sup>a</sup> | 0.3859<br>(0.121)<sup>a</sup> | 0.3857<br>(0.119)<sup>a</sup> |
| Lockdown<br>dummy (ld) | -0.1176<br>(0.228) | -0.1051<br>(0.159) | -0.0910<br>(0.130) | -0.0977<br>(0.112) | -0.0908<br>(0.100) | -0.0940<br>(0.092) |
|  | ~20200411 | ~20200412 | ~20200413 | ~20200414 | ~20200415 | ~20200416 |
| $\alpha$ | 0.4292<br>(0.054)<sup>a</sup> | 0.4289<br>(0.053)<sup>a</sup> | 0.429<br>(0.053)<sup>a</sup> | 0.4295<br>(0.052)<sup>a</sup> | 0.4303<br>(0.052)<sup>a</sup> | 0.4303<br>(0.051)<sup>a</sup> |
| A | -0.0795<br>(0.041)<sup>b</sup> | -0.0797<br>(0.041)<sup>b</sup> | -0.080<br>(0.041)<sup>b</sup> | -0.0798<br>(0.040)<sup>b</sup> | -0.0786<br>(0.039)<sup>b</sup> | -0.0786<br>(0.039)<sup>b</sup> |
| B | 0.3847<br>(0.118)<sup>a</sup> | 0.3850<br>(0.117)<sup>a</sup> | 0.386<br>(0.116)<sup>a</sup> | 0.3858<br>(0.114)<sup>a</sup> | 0.3829<br>(0.113)<sup>a</sup> | 0.3827<br>(0.111)<sup>a</sup> |
| Lockdown<br>dummy (ld) | -0.1141<br>(0.085)<sup>c</sup> | -0.1198<br>(0.080)<sup>c</sup> | -0.132<br>(0.076)<sup>b</sup> | -0.1298<br>(0.072)<sup>b</sup> | -0.1163<br>(0.069)<sup>b</sup> | -0.1140<br>(0.066)<sup>b</sup> |
|  | ~20200417 | ~20200418 | ~20200419 | ~20200420 |  |  |
| $\alpha$ | 0.4303<br>(0.051)<sup>a</sup> | 0.4308<br>(0.050)<sup>a</sup> | 0.4307<br>(0.050)<sup>a</sup> | 0.4311<br>(0.049)<sup>a</sup> | | |
| A | -0.0786<br>(0.038)<sup>b</sup> | -0.0785<br>(0.038)<sup>b</sup> | -0.0792<br>(0.038)<sup>b</sup> | -0.0792<br>(0.037)<sup>b</sup> |  |  |
| B | 0.3826<br>(0.110)<sup>a</sup> | 0.3829<br>(0.109)<sup>a</sup> | 0.3853<br>(0.108)<sup>a</sup> | 0.3858<br>(0.106)<sup>a</sup> |  |  |
| Lockdown<br>dummy (ld) | -0.1123<br>(0.063)<sup>b</sup> | -0.1178<br>(0.061)<sup>b</sup> | -0.1212<br>(0.059)<sup>b</sup> | -0.1226<br>(0.057)<sup>b</sup> |  |  |

Note: a, b, and c represent the significance levels of 1%, 5%, and 10%, respectively; the numbers in the parentheses are standard errors; one-sided test was used for testing the null hypothesis that a parameter was equal to zero.

**Figure 1.**
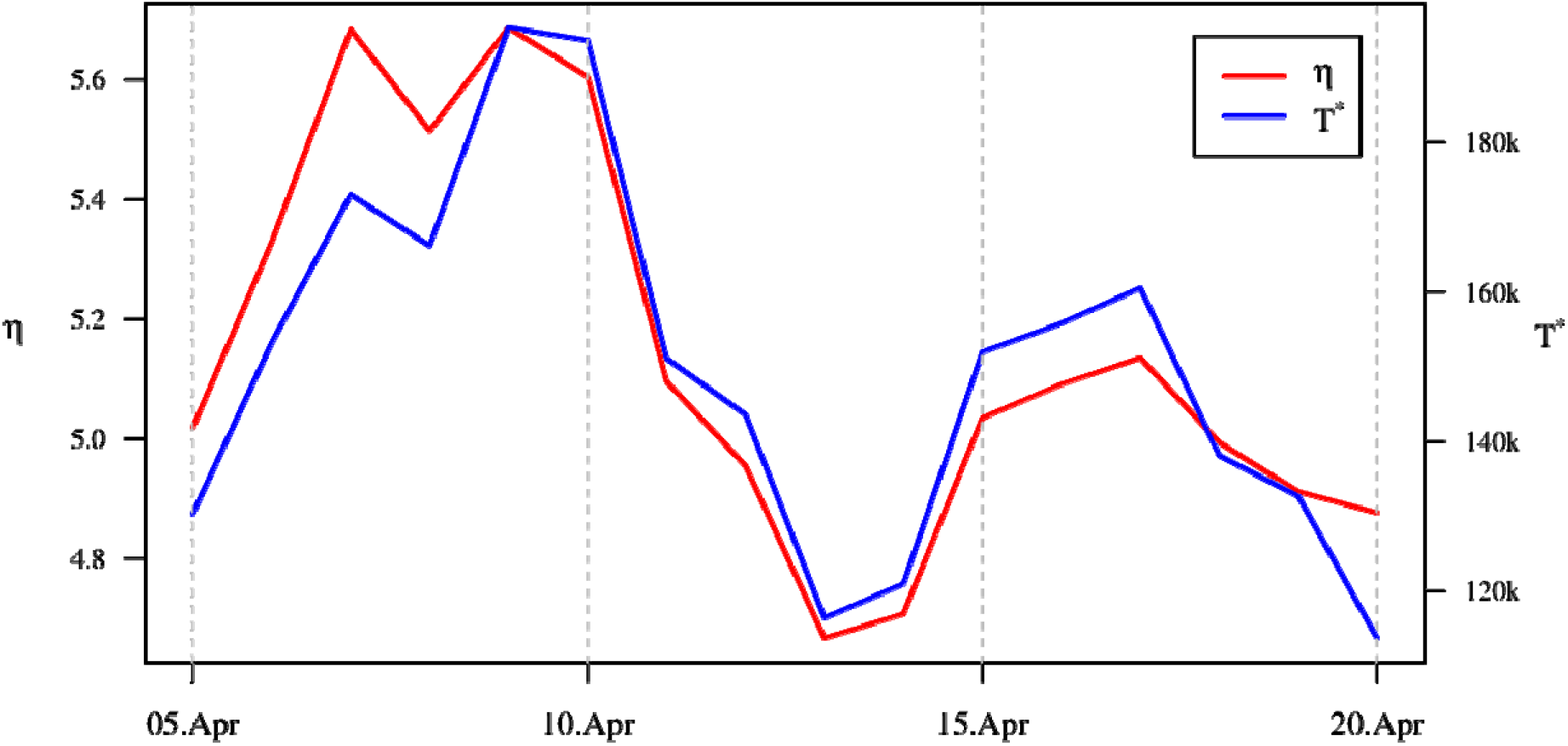
Estimation results of *η* and *T*^*^.

Figure 2 shows the estimation results of *ε* and 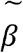 ’. Considering the structural change that takes effect after the lockdown order, we denote the new spread coefficient as 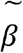 ’ to distinguish it from the original 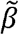. The estimates of *ε* and 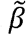 ’ were substantially stable with negligible variability. The mean value of *ε* is 2.34 and that of 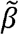 ’ is 0.478.

**Figure 2.**
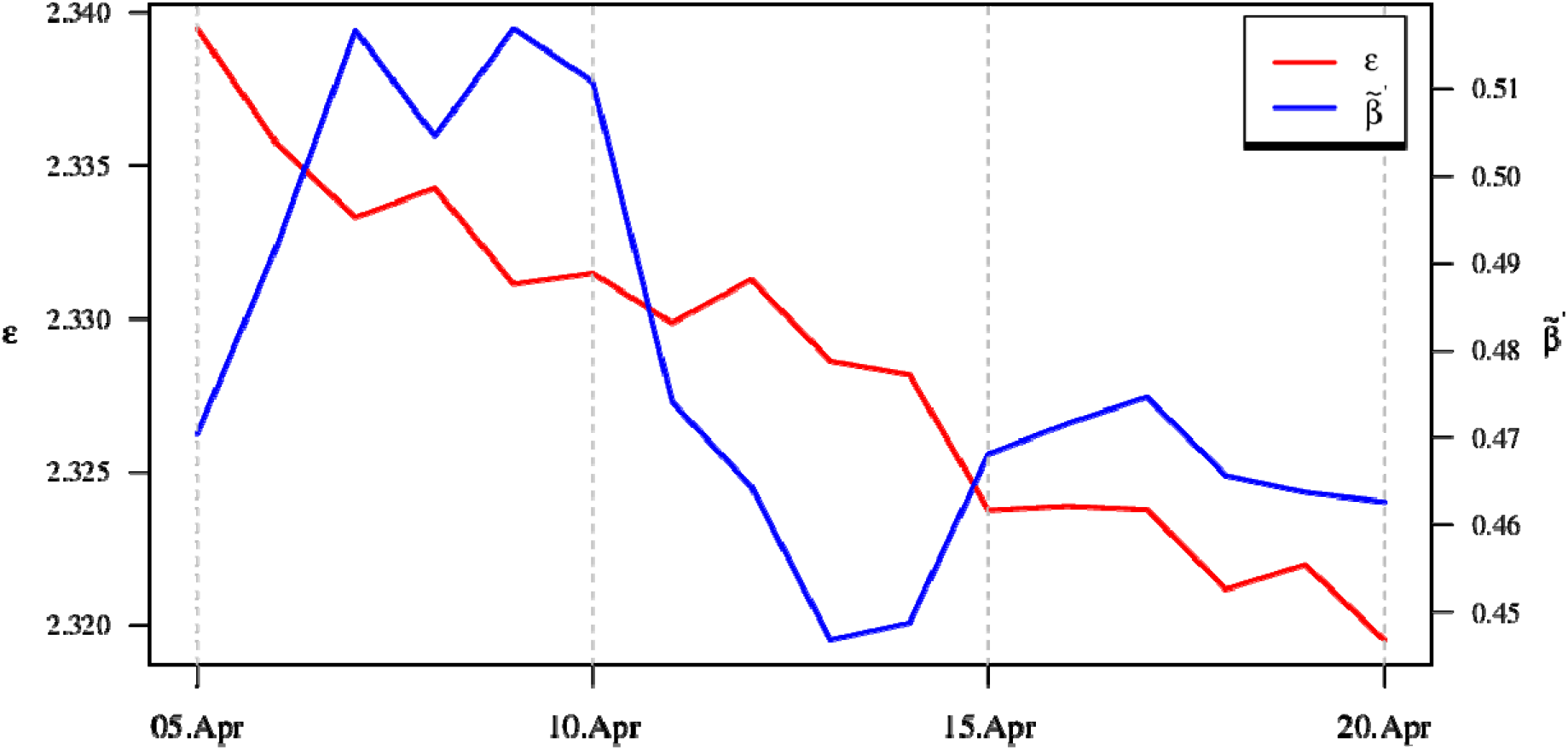
Estimation results of *ε* and 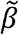 ’.

**Figure 3.**
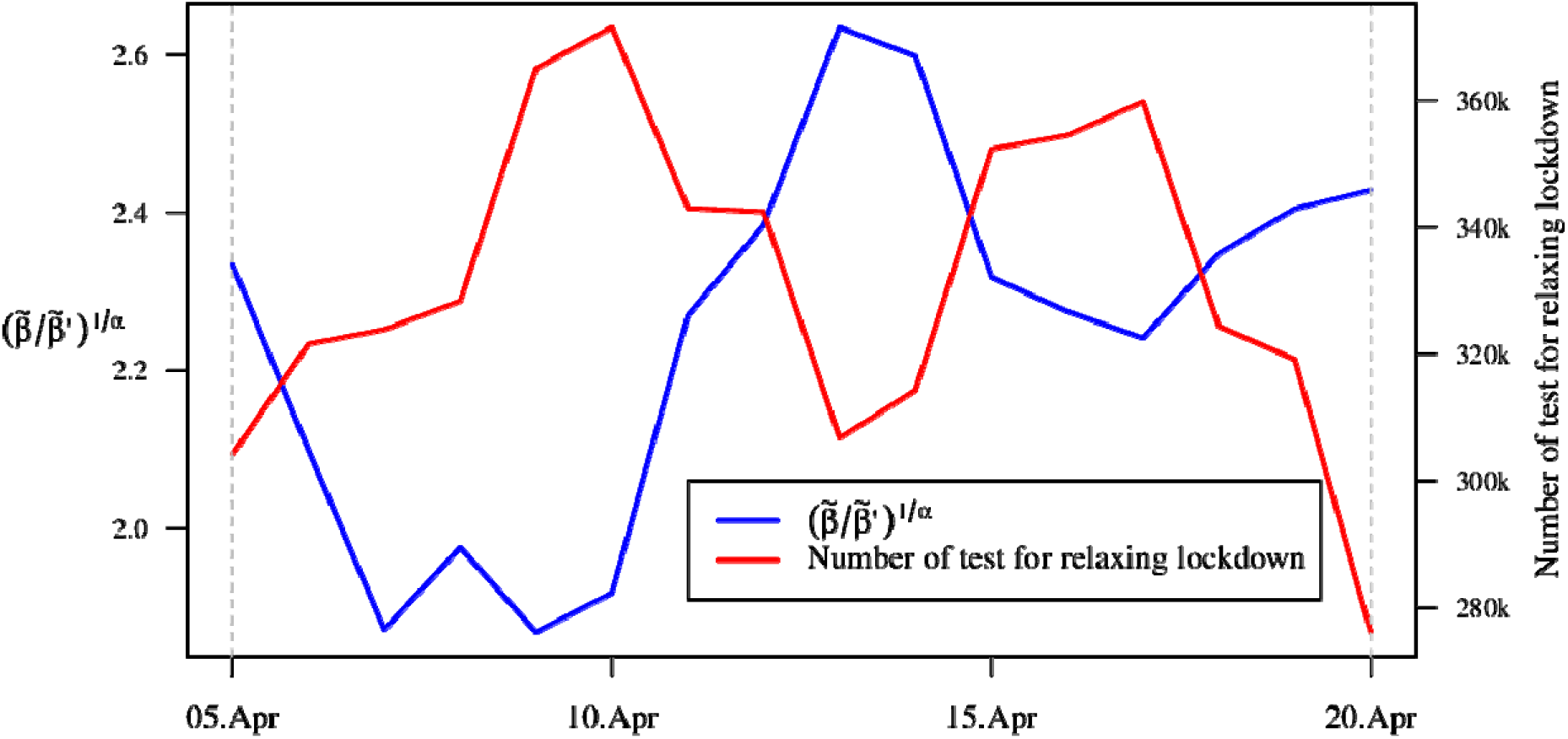
Number of tests required for relaxing lockdown.

## Discussion

For decision makers, an important question is “How many tests are required to prevent the spread when the lockdown order is relaxed?” We estimated that if the current lockdown is relaxed, the number of tests required to stop the further spread of COVID-19 would be at least 2.25 times the number of tests conducted under lockdown. In other words, if the lockdown order was relaxed on April 20, 280,000–360,000 people have been daily tested to prevent the further spread of COVID-19 in the U.S.

Although both the U.S. and South Korea confirmed their first COVID-19 cases on January 20, 2020, the number of infections in the U.S. has exceeded 2 million, whereas South Korea has flattened its epidemic curve since April (4, 5). One of the key features behind the flattening of the curve in Korea is that it adopted early measures including large-scale testing and contact tracing that significantly suppressed the outbreak (6).

The limitations of our model and estimation, such as strong mathematical assumptions and the endogeneity of the number of tests, would be interesting topics for follow-up studies. Furthermore, some of subsequent studies may extend our model to complete epidemic models by including hospitalization and recovery. Despite this, our study provides quantified evidence to the public policy regarding resource allocation for suppressing the spread of COVID-19. In the circumstance that many countries have gradually relaxed social distancing only with the expectation for herd immunity to be built-up, this study is believed to help policy-makers develop policy tools for the relaxation through the increase in the number of testing (7). Although the optimal number of testing is different across countries, the figures estimated in this study are expected to pave the road for suppressing the spread of COVID-19 (8).

In summary, we present a trade-off relationship between social distancing and mass testing, suggesting the additional mass-testing of at least 2.25 times in the case of the relaxation of social-distancing norms in the U.S. These approaches can disseminate the information regarding the decisions made by public health bodies of other countries, especially those planning to relax lockdown norms, thereby enabling both the selective adoption of policies that have proved effective in curtailing the spread of COVID-19 and additional mass testing.

## Data Availability

All the data can be retrieved from the authors.

