## Supplementary material for "Number of tests required to flatten the curve of coronavirus disease-2019": Supp

### **Model equations**

We assume that a group of people who are suspected of COVID-19 infection and are therefore being tested (the group includes both uninfected and infected people) has an exponential distribution according to symptoms. The infected people among the suspected group also have an exponential distribution in this setting. Let us denote the symptom as *x*, intercept of the distribution function of the suspected people ($\tilde{B}$) as $k_{B}$, and intercept of the distribution function of the infected people ($\tilde{I}$**)** as $k_{C}$. Thus, we obtain the following:

$\tilde{\boldsymbol{I}}\boldsymbol{=}\boldsymbol{k}_{\boldsymbol{C}}\boldsymbol{e}^{\boldsymbol{-}\boldsymbol{\alpha}_{\boldsymbol{C}}\boldsymbol{x}}$	(S1)

$\tilde{\boldsymbol{B}}\boldsymbol{=}\boldsymbol{k}_{\boldsymbol{B}}\boldsymbol{e}^{\boldsymbol{-}\boldsymbol{\alpha}_{\boldsymbol{B}}\boldsymbol{x}}$ , (S2)

where $\alpha_{C}$ and $\alpha_{B}$ denote the parameters of the two exponential distribution functions $\tilde{I}\mathrm{and} \tilde{B}$, respectively. We assume that the ratio of the number of confirmed cases to the number of tests is unity when the severity of symptoms is greater than a threshold (i.e., $x\geq\bar{x})$, where $\bar{x}$ satisfies the relation $\frac{k_{C}}{\alpha_{C}}e^{-\alpha_{C}\bar{x}}\boldsymbol{=}\frac{k_{B}}{\alpha_{B}}e^{-\alpha_{B}\bar{x}}$**.** Integrating $\tilde{I}$ over zero to infinity yields the number of infected cases per population as follows:

$\boldsymbol{I=}\boldsymbol{k}_{\boldsymbol{C}}\boldsymbol{/}\boldsymbol{\alpha}_{\boldsymbol{C}}$. (S3)

We assume that the tests are performed on suspected individuals whose symptoms range from the most severe and to as weak as $\delta$. Accordingly, the following equations are obtained:

$\boldsymbol{C=}\frac{\boldsymbol{k}_{\boldsymbol{C}}}{\boldsymbol{\alpha}_{\boldsymbol{C}}}\boldsymbol{e}^{\boldsymbol{-}\boldsymbol{\alpha}_{\boldsymbol{C}}\boldsymbol{\delta}}$	(S4)

$\boldsymbol{T=}\frac{\boldsymbol{k}_{\boldsymbol{B}}}{\boldsymbol{\alpha}_{\boldsymbol{B}}}\boldsymbol{e}^{\boldsymbol{-}\boldsymbol{\alpha}_{\boldsymbol{B}}\boldsymbol{\delta}}$, (S5)

where *C* is obtained by integrating $\tilde{I}$ over the region where the test is performed (*i.e.*, $x>\delta)$, and the number of tests per population, *T,* is obtained by integrating $\tilde{B}$ over the same region.

Furthermore, setting the coefficient of proportionality between $\frac{k_{C}}{\alpha_{C}}$ and $\frac{k_{B}}{\alpha_{B}}$ as $\mu$, we obtain the following:

$\boldsymbol{C=I}\boldsymbol{e}^{\boldsymbol{-}\boldsymbol{\alpha}_{\boldsymbol{C}}\boldsymbol{\delta}}$	(S6)

$\boldsymbol{T=}\frac{\boldsymbol{I}}{\boldsymbol{\mu}}\boldsymbol{e}^{\boldsymbol{-}\boldsymbol{\alpha}_{\boldsymbol{B}}\boldsymbol{\delta}}$. (S7)

Substituting equation (S6) into equation (S7) to eliminate $\delta$, we obtain the following:

$\boldsymbol{C=}\boldsymbol{(\mu T)}^{\boldsymbol{\alpha}}\boldsymbol{I}^{\boldsymbol{1-\alpha}}$. (S8)

Equation (S8) is of the form of the Cobb–Douglas function, which is frequently used in economics. Substituting equation (S8) into equation (1) in the main article, $\frac{dI}{dt}=\beta IS-\gamma I-C$, we obtain the following:

$\frac{\boldsymbol{dI}}{\boldsymbol{dt}}\boldsymbol{=\beta IS-\gamma I-}\boldsymbol{(\mu T)}^{\boldsymbol{\alpha}}\boldsymbol{I}^{\boldsymbol{1-\alpha}}$. (S9)

This model aims to describe the situation in which the spread is prevented via human efforts, including testing and quarantining, during the early phase of an epidemic. Therefore, assuming $S=1$, equation (S9) can be rewritten as follows:

$\frac{\boldsymbol{d}\ln\boldsymbol{I}}{\boldsymbol{dt}}\boldsymbol{=}\tilde{\boldsymbol{\beta}}\boldsymbol{-}{\boldsymbol{(}\frac{\boldsymbol{\mu T}}{\boldsymbol{I}}\boldsymbol{)}}^{\boldsymbol{\alpha}}$, (S10)

where $\tilde{\beta}=\beta-\gamma$. According to equation (S10), the growth rate of the number of infected cases is determined by i) $\tilde{\beta}$, and ii) the ratio of the number of tests to the number of infected cases, *T*/*I*. If the infectious disease continuously spreads (i.e., $\tilde{\beta}>0$), the number of tests required to deaccelerate the increase in the number of infected cases can be calculated by applying the condition that the right-hand side of equation (S10) should be less than or equal to zero. Accordingly, the minimum number of tests required to prevent the increase in the number of infected cases, *T*^*^, is obtained as follows:

$T^{*}=\frac{\tilde{\beta}^{1/\alpha}}{\mu}I$. (S11)

Notably, the number of infected cases per population, *I*, cannot be directly measured. Substituting *I* with *C* from equation (S8), the minimum number of tests required to prevent the increase in the number of infected cases per population is obtained as follows:

$T^{*}=\frac{\tilde{\beta}^{\frac{1-\alpha}{\alpha}}}{\mu}C=\eta C$. (S12)

Notably, the information regarding the number of confirmed cases yields the minimum number of tests required to prevent the spread of the disease (i.e., $\frac{d\ln I}{dt}\boldsymbol{\leq0}$). The implication of equation (S12) is as follows: To stop the spread of the infectious disease, the number of tests should be at least $\eta$ times the number of cases confirmed.

**Model fitting**

Our model provides implications regarding the change in the minimum number of tests if the societal situations, such as social distancing and lockdown, change. Once the spread of the infectious disease subsides, relaxing social distancing or lockdown policies may reduce the adverse economic and social effects. However, relaxing social distancing or lockdown policies increases $\tilde{\beta}$, which in turn increases the spread. The key to preventing the spread is to determine the number of tests required. Assuming that other state variables remain unchanged, once the lockdown is suddenly relaxed, the minimum number of tests required to ensure that $\frac{d\ln I}{dt}\boldsymbol{\leq0}$ can be estimated using the elasticity of $\tilde{\beta}$ with respect to $T^{*}$ by employing equation (S11) as follows:

$\varepsilon_{\beta}^{T}=\frac{1}{\alpha}$ . (S13)

Taking logarithm on both the sides of equation (S8) and differentiating with respect to time *t*, we obtain the following:

$\frac{\boldsymbol{d}\ln\boldsymbol{C}}{\boldsymbol{dt}}\boldsymbol{=\alpha}\frac{\boldsymbol{d}\ln\boldsymbol{T}}{\boldsymbol{dt}}\boldsymbol{+(1-\alpha)}\frac{\boldsymbol{d}\ln\boldsymbol{I}}{\boldsymbol{dt}}$. (S14)

Substituting equation (S10) into equation (S14), we obtain the following differential equation for the confirmed case:

$\frac{d\ln C}{dt}=\alpha\frac{d\ln T}{dt}+(1-\alpha)\left[ \tilde{\beta}-\left( \frac{\mu T}{I} \right)^{\alpha} \right]$. (S15)

Because ${(\frac{\mu T}{I})}^{\alpha}={(\frac{C}{\mu T})}^{\rho}$ from equation (S8), we express the epidemic model in terms of confirmed cases and testing by using equation (S15) as follows:

$\frac{\boldsymbol{d}\ln\boldsymbol{C}}{\boldsymbol{dt}}\boldsymbol{=\alpha}\frac{\boldsymbol{d}\ln\boldsymbol{T}}{\boldsymbol{dt}}\boldsymbol{+}\left( \boldsymbol{1-\alpha} \right)\left[ \tilde{\boldsymbol{\beta}}\boldsymbol{-}\left( \frac{\boldsymbol{C}}{\boldsymbol{\mu T}} \right)^{\boldsymbol{\rho}} \right]\boldsymbol{,}$	(S16)

where $\rho=-\frac{\alpha}{1-\alpha}$.

Next, we present a method for estimating the parameters of the model for empirical investigation by using actual data. Additionally, the exogenous variable, *T*, is determined *ex ante* on the basis of the government policy. Because the data are discrete and compiled daily, equation (S16) can be rewritten as follows:

$\boldsymbol{\Delta}\ln\boldsymbol{C}\boldsymbol{=\alpha\Delta}\ln\boldsymbol{T}\boldsymbol{+A}\left( \frac{\boldsymbol{C}}{\boldsymbol{T}} \right)^{\boldsymbol{-}\frac{\boldsymbol{\alpha}}{\boldsymbol{1-\alpha}}}\boldsymbol{+B}$, (S17)

where $A=-\left( 1-\alpha\right)\mu^{-\rho}=-\left( 1-\alpha\right)\mu^{\frac{\alpha}{1-\alpha}}$, and $B=\left( 1-\alpha\right)\tilde{\beta}$. Additionally, $\boldsymbol{\Delta}\ln\boldsymbol{C}$ denotes the daily rate of change in the number of confirmed cases per population ; $\boldsymbol{\Delta}\ln\boldsymbol{T}$ denotes the daily rate of change in the number of people who are tested for COVID-19 per population; iii) A and B are constants. In equation (S17), *B* is related to the rate of infection and is affected by social distancing or lockdown policies.

**Data source and estimation**

For the empirical investigation, we compiled the daily number of tests and confirmed cases from the COVID tracking project (<https://covidtracking.com>) for the period ranging from February 28 to April 20, 2020. We assumed that the effect of the lockdown policy was reflected in the data. Therefore, we examined the validity of such assumptions via a structural change model by adding a dummy variable for the lockdown policy, along with a non-structural model. We used a nonlinear regression method by adding the lockdown-policy dummy variable to the right-hand side of equation (S17). The nls () function in R-3.6.1 was used for estimating the parameters of equation (S17) .

The primary objective of estimating equation (S17) was to determine the minimum number of tests,
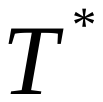
, required to reduce the number of infected cases. The estimated value of
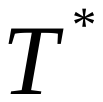
 helps policymakers decide the number of tests to be conducted in the near future. We fit the model to the daily data from February 28 to a specific date. For example, using the information regarding the number of daily infected cases collected from February 28 to April 5, the
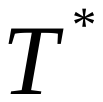
value that should be applied on April 6 can be estimated; similarly, using the information regarding the number of daily infected cases collected from February 28 to April 6, the
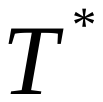
value that should be applied on April 7 can be estimated, and so on.

We must consider a certain time span to consider the time taken for the lockdown order to become effective. For example, although a lockdown order was announced by the federal government of the U.S. on March 21, the effect of the lockdown order was assumed to manifest from April 5, considering the time taken by the U.S. public to accept and implement the lockdown order.

The estimation results of the number of confirmed cases are depicted in Figure 1. From March 1 to March 16, 2020, the estimated number of confirmed cases is significantly close to the actual number of confirmed cases. Although some differences are observed between the estimated and actual values from March 16 to April 5, the surge in the number of confirmed cases is appropriately estimated using the proposed model. Although some differences do exist between the estimated and actual measurements from April 5 onward, our estimation results indicate that our proposed model can accurately capture a steady pattern of the number of actual confirmed cases with lockdown and mass testing, and this is the main strength of our mathematical model.

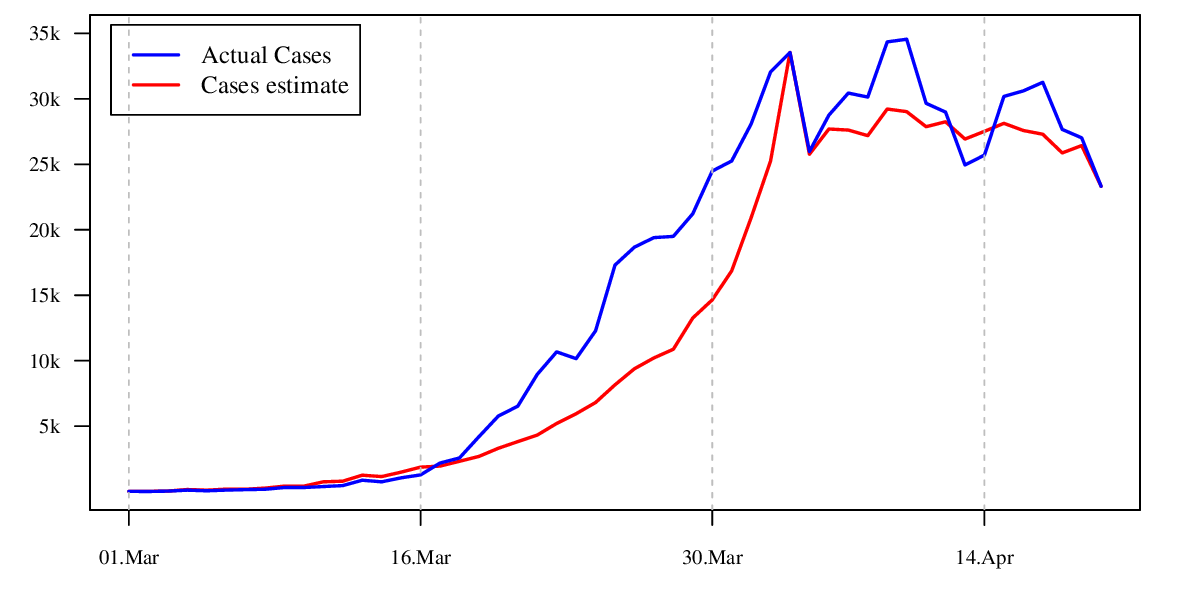

Figure 1. Number of actual and estimated cases.
